# Multivariate prediction of temper outbursts in youth enriched for irritability using Ecological Momentary Assessment data

**DOI:** 10.1101/2023.07.14.23292689

**Authors:** Dipta Saha, Reut Naim, Francisco Pereira, Melissa A. Brotman, Charles Y. Zheng

## Abstract

Irritability and temper outbursts are among the most common reasons youth are referred for psychiatric assessment and care. Identifying clinical variables (e.g. momentary anxiety) that precede the onset of temper outbursts would provide valuable clinical utility. Here, we provide the rationale for a study to test the performance of classifiers trained to predict temper outbursts in a group of clinically-referred youth, in a home setting, enriched for symptoms of irritability and temper outbursts. Using observational data--digital based event sampling from previous Ecological Momentary Assessment data, we demonstrate promising results in our ability to predict the presence of a temper outburst based on clinical responses (e.g., whether the participant is grouchy, hungry, happy, sad, anxious, tired, etc.) prior to the emotional event, as well as external features (e.g., time of day, day of week). In exploratory analyses of existing data, consisting of n=57 subjects with a total of 1296 time points, we evaluate the feasibility of using a logistic regression-based classifier and a random-forest based classifier for predicting the temper outburst prospectively. In order to more rigorously assess these classifiers, we propose the collection of a large confirmatory set, consisting of at least an additional 20 subjects with an expected total of 400 time points, and will perform confirmatory analyses of the precision and recall of several classifiers for predicting temper outbursts. This work provides the foundation for the identification of features predictive of risk and future development of novel mobile-device-based interventions in youth affected with severe and impairing psychopathology.

## Introduction

Irritability and temper outbursts are one of the most common reasons for child mental health referrals (Evans et al. 2022, Peterson et al. 1996, Brotman et al. 2017). The use of real-time, *in vivo* metrics using smartphones and other wireless devices provides the possibility of obtaining naturalistically situated information on the psychological state of children with symptoms of irritability (i.e. disruptive mood dysregulation disorder (DMDD), oppositional defiant disorder (ODD), sub-threshold symptoms of DMDD (sub-threshold DMDD)) by means of digital-based phenotyping using ecological momentary assessment (EMA) (Naim et al. 2021, Naim et al. 2022, Wen et al. 2017). EMA data has the potential to provide the clinician with valuable insight into longitudinally-presenting dynamics of irritability and other related clinical symptoms. Here, we investigate, in a clinically enriched observational study, the degree to which dynamic phenotyping via EMA enables the prospective prediction of temper outbursts. Previous work has demonstrated the feasibility of predicting symptomatic behavior from psychological traits (Arria et al. 2009, Dahlen et al. 2005). Models which succeed at predicting temper outbursts would be valuable not only because of the possibility of revealing predictive relationships between irritability and other time-varying metrics (including symptoms, time of day, and hunger), but also because of the possibility of using such models as an early warning system within a device-assisted intervention program. In this registered protocol, we describe the development of a classification model on existing data, and our planned validation of the model on data that we plan to collect.

## Study Information

### Hypotheses

In an ecological momentary assessment digital platform that queries real-time symptom metrics three times a day, and in a model that is shared across subjects, we hypothesize that it is possible to prospectively predict a temper outburst in the next ecological momentary assessment (EMA) rating using the following measurements:

-- The subject’s age and self-reported sex assigned at birth.

-- The participant’s momentary state of feeling: tired, worried, happy, grouchy, angry, frustrated, worried, unhappy, having experienced a change in mood.

-- Whether the day of the week is Sunday, Monday, Tuesday, Wednesday, Thursday/Friday, Saturday, or Sunday (Thursday and Friday being considered as one category.)

-- Whether the time of day is evening, or before evening (morning and afternoon being considered as one category).

### Design Plan

#### Study Type

The study is an observational study, since data is collected from study subjects that are not randomly assigned to a treatment.

#### Blinding

No blinding is involved in this study. The authors had access to information that could identify participants during and after collection of data.

#### Study design

The data was collected according to a protocol from Naim et al. 2021. No participants were treated during the data collection. Quoting from Naim et al. 2021:

“Youth participants enriched for symptoms of irritability and temper outbursts and an identified parent completed a clinical evaluation visit before being enrolled in the research protocol. Participants subsequently completed a standardized EMA training session during which a research assistant familiarized the participant with the smartphone and protocol and reviewed each EMA item by guiding the participant through a practice prompt. To enhance feasibility and compliance, for each day during the upcoming 7 days of EMA, participants preselected 60-min periods during standardized time windows within which prompts would be delivered: morning/before school (6:00–9:00 a.m.), afternoon/after school (3:00–6:00 p.m.), and evening/before bedtime (7:00–10:00 p.m.). The actual prompt times were randomized within these time periods… Participants used either a personal or a study-provided smartphone…

Following the training session, participants were prompted three times per day for seven consecutive days. At each prompt, participants received a text message with a link to the website through which the items were delivered. Once the prompt was received, participants had 60 min to complete the assessment before it expired and was considered incomplete.”

This protocol was carried out from July 2018 and is currently ongoing at NIMH. Participants were recruited from the Washington, DC, metropolitan area in the context of 2 ongoing studies (ClinicalTrials.gov Identifiers NCT02531893 and NCT00025935). Recruitment strategies leveraged established relationships with local health care providers and schools and included advertising on Facebook and sending postcards to local households. Outpatients were referred to NIMH.

### Sampling Plan

#### Explanation of existing data

We split data collection into two stages. In the first stage, which was collected starting July 2018 and ending in December 2022, n=57 subjects with ODD/DMDD/sub-threshold DMDD were collected (and 3 were excluded for not having enough EMA ratings during the recording period), with a total of 1296 ratings from non-excluded subjects. We have completed the first stage, inspected the data, and fit and evaluated models to the data.

**Table 1.** Demographic characteristics and outcome statistics for subjects from existing data included in the analysis.

|  |  |
| --- | --- |
| Number of non-excluded subjects | n=54 |
| Age range | 8-18 |
| Mean age | 11.6 |
| Number of male subjects | n=34 |
| Number of female subjects | n=20 |
| Mean number of complete ratings | 24 |
| Number of subjects with at least one missing rating | n=44 |
| Mean (sd) number of temper outbursts | 0.100 (0.160) |
| Mean (sd) number of temper outbursts within males | 0.056 (0.080) |
| Mean (sd) number of temper outbursts within females | 0.175 (0.222) |

The results from the initial data analysis are as follows.

We formed a training set, stage 1a, from data collected before 2023. The validation set, stage 1b, was formed from data collected in January 2023, and was used to assess model performance. The sample size was thus determined by the size of the existing data and the need to split the data to enable unbiased inference of the classifier performance.

We considered 23 variables. We eliminated 6 variables by means of backwards selection algorithm using Bayesian comparison tests (Benavoli et al. 2017). The following 17 variables were selected.

-- The subject’s age and self-reported sex assigned at birth.

-- The participant’s momentary state of feeling: tired, worried, happy, grouchy, angry, frustrated, worried, unhappy, having experienced a change in mood.

-- Whether the day of the week is Sunday, Monday, Tuesday, Wednesday, Saturday, or

Sunday

-- Whether the time of day is evening.

We excluded the following variables from the model based on the results of variable selection.

--Whether the subject has been hungry, or giddy.

--Day of week: Thursday and Friday.

--Time of day: morning and afternoon.

In table 2, we present the model performance for regularized logistic regression.

**Table 2.** Prediction performance of temper outbursts for DMDD, sub-threshold DMDD, and ODD patients in existing dataset, using regularized logistic regression. Precision, also called positive predictive value, is defined as the fraction of positive predictions that were true positives (the fraction of times where a temper outburst was endorsed out of the times that the model predicted a temper outburst). Recall, also called sensitivity, is defined as the fraction of positive examples that were predicted to be positive (the fraction of times where the model predicted a temper outburst out of the times in which a temper outburst was endorsed). F1-score is the harmonic mean of precision and recall, given by the formula F1 = 2 * (precision * recall) / (precision + recall). Support is the total number of test set observations in each category.

|  | Precision | Recall | F1-score | Support |
| --- | --- | --- | --- | --- |
| No outburst | 0.96 | 0.75 | 0.84 | 331 |
| Temper outburst | 0.23 | 0.71 | 0.34 | 34 |

**Table 3.**
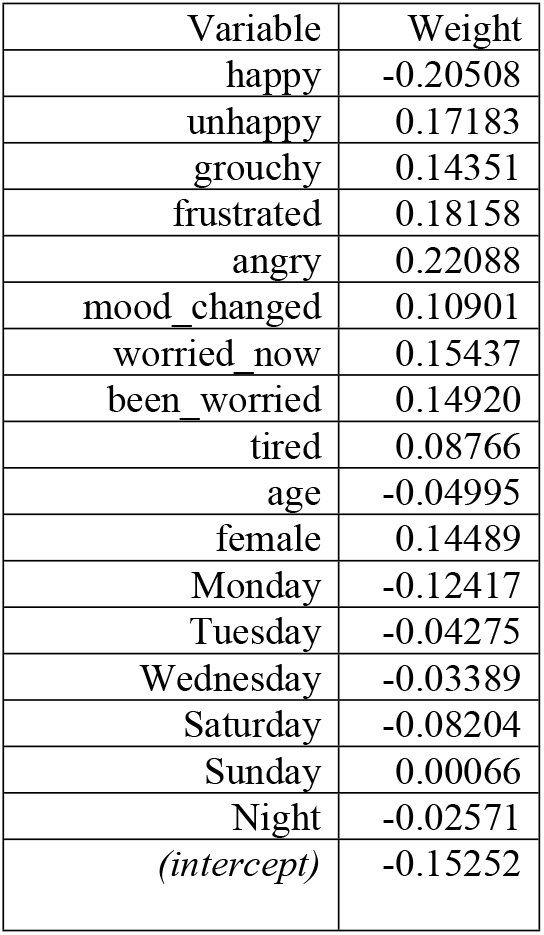
Model weights and intercept for logistic regression. A positive (negative) value indicates that a YES response is associated with increased (decreased) chance of temper outburst in the next measurement period. For questions corresponding to labels, please see table 3.

We also tried using the g-mean (Kubat and Matwin 1997) to select a probability threshold for positive prediction. The threshold chosen was 0.504, very close to the default threshold 0.5, so that results were unchanged.

The model weights for logistic regression were as follows:

#### Data Collection Procedures

Recruitment focused on youth aged 8–18 years (Mage = 11.574 years, SD = 2.249 years; 62.96% male for included subjects) meeting criteria for DMDD (n = 38), sub-threshold DMDD (n=10), or ODD (n = 6). Some of the participants also had co-occuring anxiety disorder(s) and/or attention-deficit hyperactivity disorder (ADHD).

The means of recruitment and exclusion criteria are described in Naim et al. 2021, as quoted:

“Participants were recruited via direct mailings and online advertisements. Participants were evaluated for eligibility and diagnostic status by a doctoral- or master’s-level clinician using the Schedule for Affective Disorders and Schizophrenia for School-Age Children – Present and Lifetime version (Kaufman et al. 1997). Primary diagnosis was based on the chief presenting complaint and clinician judgment of the most severely impairing diagnosis. Recruitment focused on youth whose irritability was chronic and not clearly related to another ongoing or episodic diagnosis (e.g., major depressive disorder, bipolar disorder). Exclusion criteria were: IQ < 70, assessed using the Wechsler Abbreviated Intelligence Scale (Wechsler 1999); a diagnosis of posttraumatic stress disorder, schizophrenia, neurological disorder, developmental disorder, bipolar disorder, or obsessive–compulsive disorder; a current major depressive episode; or substance abuse within 3 months of participation. Participants and their parents provided written assent and consent, respectively. Participants were compensated and offered a monetary bonus for completing≥ 75% of prompts. The study was approved by the National Institute of Mental Health Institutional Review Board.”

#### Sample size

In the first stage, we collected 12 participants with 1 week of data, and 39 participants with 2 weeks of data.

In the second stage (to be completed by approximately 2025), we will collect 20-40 participants with a total of at least 40 weeks of data across all participants, and an expected total of 400 ratings. We determined this prospective sample size based on the number of ratings needed to have sufficient positive observations given an estimated frequency of 10% for the probability of temper outburst.

#### Sample size rationale

We collect 20-40 participants with at least 40 weeks of data total, to ensure sufficient sample size and diversity to validate the prediction performance of our model.

### Variables

#### Measured variables

Here we present the details of selected variables. For details on excluded variables, see Naim et al. 2021.

- Irritability symptoms (Details for the rationale of these measures can be found in Naim et al. 2021.)
  - (a)temper outburst – “SINCE the last beep, I felt really, really angry and out of control.” (categorical yes or no)
  - (b)irritable mood – “SINCE the last beep, aside from being really, really angry and out of control, I was feeling generally grouchy or cranky.” (5-point Likert scale, 1 = none of the time; 5 = the whole time)
  - (c)frustration – “SINCE the last beep, I felt frustrated.” (5-point Likert scale, 1 = not at all; 5 = extremely)
  - (d)momentary anger – “At the time of the beep, I felt ANNOYED or ANGRY.” (5-point Likert scale, 1=not at all; 5= extremely)
- Anxiety symptoms (Details for the rationale of these measures can be found in Naim et al. 2021.)
  - (a)anxious affect – “SINCE the last beep, I felt worried or scared.” (5-point Likert scale, 1 = not at all; 5 = extremely)
  - (b)anxious avoidance – “SINCE the last beep, I avoided doing things because I felt worried or scared.” (categorical yes or no)
  - (c)momentary anxiety (Smith et al. 2022) – “At the time of the beep, I felt WORRIED or SCARED.” (5-point Likert scale, 1 = not at all; 5 = extremely)
- The participant’s momentary feeling state:
  - (a)tired -- “At the time of the beep, I felt TIRED.” (5-point Likert scale, 1 = not at all; 5 = extremely)
  - (b)worried -- “At the time of the beep, I felt WORRIED or SCARED.”
  - (c)happy -- “At the time of the beep, I felt HAPPY.”
  - (d)grouchy -- “SINCE the last beep, aside from being really, really angry and out of control, I was feeling generally grouchy or cranky.”
  - (e)unhappy -- “At the time of the beep, I felt unhappy, sad, or miserable.”
  - (f)having experienced a change in mood -- “SINCE the last beep, my mood changed a lot.” (5-point Likert scale, 1 = not at all; 5 = extremely)

### Analysis Plan

#### Statistical Models

##### Data

The full list of selected variables is as follows.

Response variable: Temper outburst (binary) Predictor variables:

1. non-time-varying:
  1. Demographic variables: age and self-reported sex assigned at birth
2. time-varying:
  1. Indicator variables from previous time point: giddy, tired, worried, happy, grouchy, angry, frustrated, worried, unhappy, change in mood.
  2. Time of day and day of week, see “Measured Variables” for details

##### Analytic approach

We use two models: logistic regression and random forest.

Time-varying predictors are included with a lag of 1, so that only information proceeding a temper outburst is used to predict the absence/presence of a temper outburst. All variables were scaled with min-max scaling to lie between 0 and 1.

##### Logistic Regression

We use logistic regression with either L1 or L2 regularization (see below for details on hyperparameter selection) and balanced class weights. Furthermore, we use Bayesian comparison of classification models (Benavoli et al. 2017) to perform backwards variable selection.

To select hyperparameters, we use stratified cross-validation with 40 splits to search over a hyperparameter grid. The number of splits was chosen to achieve a balance between having a large number of splits (which improves classifier performance) and ensuring that each split had a minimal number of temper outbursts (e.g. 1 or 2). We pick the hyperparameter combination (penalty + cost parameter) that minimizes the F2 score. The F2 score is used to give more weight to recall than to precision, as we consider it acceptable to have a high percentage of false alarms in order to achieve a high coverage of prospective temper outbursts.

We used the following grid parameters:

- “C” (cost parameter, inverse of alpha parameter): {.001,.01,.1,1,10,100,1000}
- penalty: L1, L2

We used scikit-learn (Pedregosa et al. 2011) to implement the regression.

##### Random Forest

On the same list of variables, we also consider using a balanced random forest. As with logistic regression, we used balanced class weights, and used stratified 40-fold cross-validation to pick hyperparameters. The hyperparameters are:

- Number of trees: {5,10,20,50,100,120}
- Maximum depth: {1,2,3,4,5,6,7,8}
- Split criterion: Gini or Entropy

We used imbalanced learn (Lemaître et al. 2017) to implement the random forest.

#### Transformations

The day of week was extracted from the timestamp of the raw data.

Recall that the data were collected for 7 days, 3 times a day per observation period, with some subjects completing multiple observation periods (details in sampling plan). The time of day was obtained from the column number of the raw data (as columns 1-21 are the 21 consecutive EMA rating periods, with morning, afternoon, and evening for each of 7 days).

All predictors were rescaled to lie between 0 and 1 using min-max scaling. Observations with missing entries were excluded from the analysis. Since we do not use observations with partial entries, we are reporting the missing cases on an observation-by-observation basis rather than a variable-by-variable basis.

#### Inference Criteria

We use Bayesian model comparison (Benavoli et al. 2017) to perform feature selection using the training set (the data collected in the first stage). Specifically, we use the Bayesian paired t-test with one-sided posterior probabilities.

In order to carry out backwards feature selection, we use the following procedure. Given a model (list of features), we:

1)compare (using the Bayesian paired t-test) the model to all versions of the model with one variable deleted,

2) if any model with a deleted variable has a higher than 50% probability of being better than the original model, take the model with the highest probability of being better than the original model, and repeat step 1 with that model.

This procedure stops when none of the deleted features has at least a 50% posterior probability of performing better than the current model. The current model is therefore selected for use on the test set.

On the test set to be collected, we will compute confidence intervals for the precision and recall of the model (Goutte and Gaussier 2005).

#### Exploratory Analysis

During the model development process, we examined the pairwise correlations between variables in order to identify pairs of variables that were likely to be redundant. We found that “happy” and “unhappy” were highly negatively correlated, and that “unhappy”, “grouchy” and “frustrated” were highly positively correlated, as we show in Figure 1.

**Figure 1.**
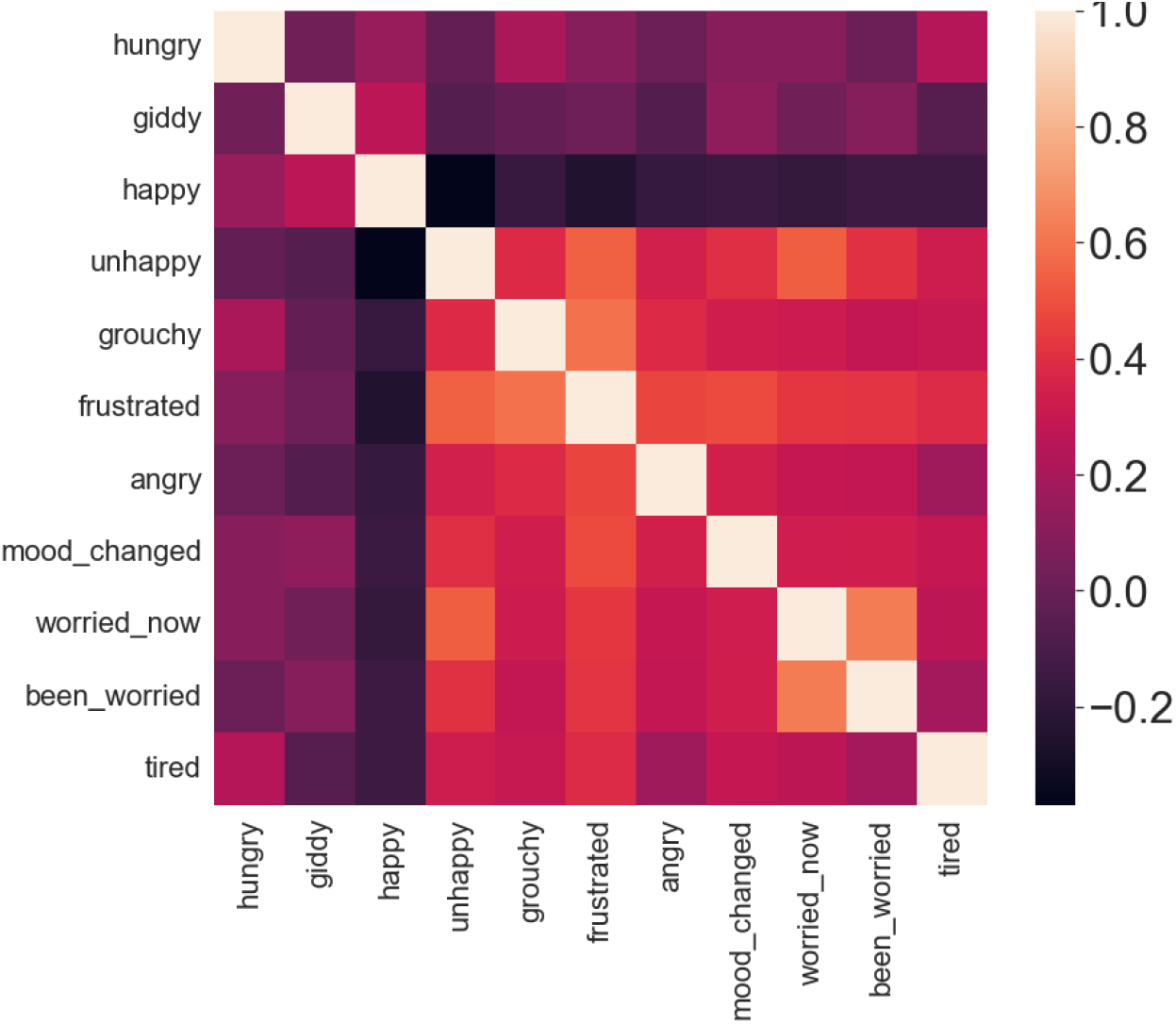
Correlations between some of the variables. We found that “happy” and “unhappy” were highly negatively correlated, and that “unhappy”, “grouchy” and “frustrated” were highly positively correlated. See table 3 for the questions corresponding to the labels.

**Table 3.** Label key for figure 1 and table 3.

| Label | Question |
| --- | --- |
| hungry | "At the time of the beep, I felt HUNGRY." |
| giddy | "AT THE TIME OF the beep, I felt much more giddy, silly, or happy than usual. " |
| happy | "At the time of the beep, I felt HAPPY." |
| unhappy | "At the time of the beep, I felt unhappy, sad, or miserable." |
| grouchy | "SINCE the last beep, aside from being really, really angry and out of control, I was feeling generally grouchy or cranky." |
| frustrated | "SINCE the last beep, I felt frustrated." |
| angry | "At the time of the beep, I felt ANNOYED or ANGRY." |
| mood changed | "SINCE the last beep, my mood changed a lot." |
| worried now | "At the time of the beep, I felt WORRIED or SCARED." |
| been worried | "SINCE the last beep, I felt worried or scared." |
| tired | "At the time of the beep, I felt TIRED." |

## Discussion

In this registered protocol, we propose a study for assessing the potential of machine-learning-based methods for enabling multivariate prediction of temper outbursts.

Identifying the extent to which temper outbursts can be predicted could be employed in future interventions for pediatric irritability, such as informing pediatric patient, parents and treating clinicians to the child’s clinical status. In future work, EMA reports could also be paired with real-time monitoring of youth’s physiology and behavior to derive a more complete phenotype of irritability and develop precise temporally-informed intervention targets.

There are two major limitations to this proposed study. Firstly, the sample will be nonrepresentative, as the study consists of individuals who choose to participate in NIMH studies. To mitigate the bias due to the nonrepresentative study population, we recruit across the community. Furthermore, some bias may be due to the fact that participation is potentially limited by factors that facilitate remote research compliance. Reasons for lack of participation include finding the task to be boring or annoying, or conflict between the task and existing activities. To increase compliance, we allowed participants to adjust the scheduling of the responses within 3-hour intervals for morning, noon, and evening, and randomized the timing of the prompts within the selected 60-minute interval for each time period. However, we recognize that the freedom to select the response period may lead to additional bias due to possible confounding between preferences for timing windows and measured behavior.

## Data Availability

Data is available upon request, in line with NIH and IRP guidelines.

